# Sleep duration as a predictor and prognosticator for cancer survivors in the community

**DOI:** 10.1101/2025.06.17.25329818

**Authors:** Jun Song, Chaoqun Liang, Chao Zhang, Long Liu, Junwei Yu, Nan Cheng, Yun Xiao, Han Wu, Jianming Yang

## Abstract

**Objective:** Sleep and cancer are intricately interconnected. We used a nationally representative sample to study the effect of sleep duration on cancer incidence and prognosis, aiming to propose an optimal sleep duration and generalize it to community populations.

**Methods:** Data from participants aged ≥18 years in the National Health and Nutrition Examination Survey (NHANES, 2005-2018) were analyzed (n=37,338). Self-reported sleep duration was analyzed alongside mortality data from the National Death Index as of 12/31/2019. Primary outcomes included all-cause mortality, cancer-specific mortality, and non-cancer mortality. Odds ratios (OR), hazard ratios (HR), and corresponding 95% confidence intervals (CI) were calculated using logistic regression and Cox proportional hazards regression, respectively. Restricted cubic spline (RCS) analysis was used to explore potential non-linear associations.

**Results:** The prevalence of cancer was 10.12%. After adjusting for confounders, >7 hours of sleep was strongly associated with an increased OR for cancer incidence (1.18 (1.04,1.33) for (7-8] and 1.54 (1.28,1.86) for (8-14.5]). During a median follow-up of 6.1 years, 341 cancer-related deaths and 661 non-cancer-related deaths occurred. fully adjusted models, compared with 7 hours of sleep, >7 hours of sleep significantly increased the HR of all-cause mortality (1.40 (1.06,1.84) for (7-8] and 1.90 (1.42,2.54) for (8-14.5]); cancer mortality (1.20 (0.77,1.87) for (7-8] and 1.70 (1.05,2.73) for (8- 14.5], 1.70 (1.05,2.73)) and non-cancer mortality (1.50 (1.07,2.12) for (7-8], 2.03 (1.42,2.90) for (8-14.5]). Stratified analysis, RCS analysis and sensitivity analysis yielded consistent results.

**Conclusions:** This study suggests that the optimal sleep duration should be 7 hours, as excessive sleep may increase cancer risk and mortality risk in cancer survivors. Our findings should be useful for health education and promotion in primary care and clinical practice.

## Background

Cancer is one of the leading causes of death globally and places an enormous economic burden on public health systems^1,2^. The number of cancer survivors worldwide is growing rapidly as treatments continue to improve, with 1.95 million new cancer cases and 600,000 cancer-related deaths projected for 2023 in the United States^3^. Cancer survivors face greater physical and mental health risks in the years following treatment. Notably, 50% of cancer survivors report sleep problems, a rate about three times higher than that of the general population^4^.

Sleep is essential for maintaining optimal physiological processes. Both short and prolonged sleep durations are considered important behavioral risk factors associated with multiple chronic adverse health outcomes, such as obesity, diabetes, and depression^5–8^. Emerging evidence suggests that sleep may be intricately related to cancer development^9^. First, cancer survivors may be at an increased risk for sleep problems during diagnosis, treatment, and ongoing survival. Cancer treatment measures, such as chemotherapy and radiotherapy, can cause or exacerbate sleep disorders^10^. Various preclinical studies are actively exploring the relationship between sleep and cancer development. Interestingly, recent studies have shown that cancer cells can spread more aggressively during sleep, suggesting that sleep may accelerate cancer progression^11,12^. Therefore, it is particularly important to identify the relationship between sleep duration and cancer development.

Compared to other sleep disorders, sleep duration is an easy-to-monitor and modifiable risk factor. Identifying its role in disease prediction and prevention in the general population would be of great public health importance. To our knowledge, there is a lack of comprehensive analyses on the association between sleep duration and cancer survival in community-based populations using nationally representative cohorts. Therefore, this study utilized data from the 2005-2018 National Health and Nutrition Examination Survey (NHANES) to examine the association between sleep duration and cancer prevalence in the general population. Additionally, this study analyzed the relationship between sleep duration and mortality among cancer survivors. The aim is to provide a simple, modifiable, and widely applicable predictor to identify cancer patients in the general population and to offer guidance on prognosis for cancer survivors.

## Methods

### Study population

This study included a representative sample from NHANES, a nationally representative survey conducted by the National Center for Health Statistics to assess the nutritional and health status of the U.S. non-institutionalized population^13^. Continuous NHANES is a biennial survey series that utilizes a complex, multistage probability sampling design with oversampling of various subpopulations to improve estimation accuracy. All NHANES protocols were approved by the Ethics Review Board of the National Center for Health Statistics of the Centers for Disease Control and Prevention, and all participants provided fully informed written consent. Each participant was invited to participate in a face-to-face interview, a series of physical examinations, and laboratory tests in a mobile screening facility.

To ensure the reliability and integrity of the data, the NHANES questionnaire data collection process follows standardized and tightly controlled procedures. A key role is played by the Computer Assisted Personal Interview (CAPI) system, which is programmed with built-in consistency checks to minimize data entry errors. In addition, the CAPI system includes an online help screen that provides valuable assistance to interviewers in accurately defining key terms in the questionnaire. This rigorous quality assurance and control framework underscores our commitment to maintaining high standards of data quality throughout NHANES, which enhances the credibility and robustness of our research findings.

This study analyzed NHANES data spanning from 2005 to 2018. Initially, participants under 18 years old and those lacking assessment data on cancer history were excluded. Subsequently, individuals without sleep duration assessment, pregnant participants, and those lacking weighting or follow-up information were also excluded. Informed consent and institutional review board approval were not required for this study as the NHANES public dataset used did not include personally identifiable information. This cohort study adhered to the Strengthening the Reporting of Observational Studies in Epidemiology (STROBE) guidelines for reporting.

### Sleep duration assessment

Participants self-reported the sleep duration on a typical workday or weekday. In the 2005-2014 cycle, sleep duration was created from a question posed to NHANES participants about the amount of sleep participants get on a daily basis, "how much sleep do you get (hours)?". In the 2015-2016 cycle, sleep duration was created from a question, "sleep hours". Ideally, 6-8 hours may be beneficial^14,15^, however 7 hours of sleep may be optimal^16,17^. Therefore, sleep duration is categorized as [1-6), [6-7), 7, (7-8], and (8-14.5] hours.

### Cancer assessment

The NHANES study collected information about cancer history through a self-administered questionnaire. Participants were considered cancer survivors if they met the following two criteria: 1. Those who answered "yes" to the question "Ever told you had cancer or malignancy?"; 2. Have you ever had the question "What kind of cancer? ". Participants who answered "no" to either question were used as a control group^18^.

### Mortality ascertainment

The primary outcomes of this study encompassed all-cause mortality, cancer-specific mortality, and non-cancer mortality. Survival status for the follow-up cohort was determined using the NHANES Public Use Linked Mortality file, which was matched with the National Death Index (NDI) data as of December 31, 2019. Causes of death were classified according to the International Statistical Classification of Diseases and Related Health Problems, Tenth Revision (ICD-10). All-cause mortality was defined as deaths from any cause, while cancer-specific mortality specifically referred to deaths categorized under ICD-10 codes C00-C97. Deaths from other causes were classified as non-cancer mortality. The follow-up period extended from the date of the interview to the date of death, with the final follow-up date set as December 31, 2019, for event-free individuals.

### Covariates

Covariates were selected based on prior literature and substantive reasoning. Baseline data on study participants were gathered through questionnaires and physical measurements. Self-reported variables included age (in years), gender (male or female), education level (below high school, high school, or above high school), and race/ethnicity (non-Hispanic white, non-Hispanic black, Mexican American, or other race). Body mass index (BMI) was calculated from NHANES measurements and categorized as <25.0, 25.0-29.9, or ≥30.0 kg/m². Poverty status was assessed using the poverty income ratio (PIR), calculated as household income divided by the poverty line based on household size according to U.S. Department of Health and Human Services guidelines. PIR categories included ≤1.0, 1.1-3.0, and >3.0^19^. Individuals who smoked fewer than 100 cigarettes in their lifetime were categorized as never smokers; former-smokers were defined as individuals who smoked more than 100 cigarettes in their lifetime but subsequently quit; current smokers were defined as individuals who currently smoke^20^. Drinking status was categorized as follows: non-drinkers (individuals who did not report drinking in the past 12 months), low to moderate drinkers (individuals who consumed <3 drinks per day for men, <2 drinks per day for women, or <5 episodes of binge drinking in the past 30 days), or heavy drinkers (individuals who consumed ≥4 drinks per day for men, ≥3 drinks per day for women, or ≥5 episodes of binge drinking in the past 30 days)^21^. Diabetes, hypertension, and hyperlipidemia were self-reported by participants diagnosed by a physician or other health professional. The severity of depressive symptoms over a 2-week period was assessed using the Patient Health Questionnaire-9 (PHQ-9), which employs a scale ranging from 0 to 27. Individuals scoring ≥10 were classified as having major depressive disorder; this threshold is associated with a sensitivity and specificity of 88%^22^. Leisure-time physical activity (LTPA) was evaluated using the Global Physical Activity Questionnaire (GPAQ).

Total LTPA was calculated as the sum of minutes spent in moderate-intensity recreational activities plus twice the minutes spent in vigorous-intensity recreational activities^23^. According to the 2018 Physical Activity Guidelines for Americans, participants were classified into three categories based on their LTPA levels from the previous week: inactive (no LTPA), insufficiently active (LTPA > 0 min/week but less than 150 min/week), and sufficiently active (LTPA ≥ 150 min/week)^24^.

### Statistical analysis

According to NCHS(National Center for Health Statistics) analytic guidelines, all statistical analyses using continuous NHANES data must incorporate a complex survey design to ensure estimates are representative of the U.S. noninstitutionalized civilian population. This includes using sample weights and accounting for geographic clustering indicators such as primary sampling units and strata^25^.

Continuous variables were reported as weighted means (standard error, SE), whereas categorical variables were presented as counts (weighted frequencies). Logistic regression analyses were employed to compute adjusted ORs and 95%Cis to assess associations between sleep duration and cancer incidence. Cox proportional hazard models were utilized to calculate adjusted HRs and 95% CIs for all-cause and cause-specific mortality among cancer survivors, taking sleep duration into account.

The stratified analyses were conducted based on age (<45, ≥45 years)^26,27^, gender (male, female), race (white, other), BMI (<30, ≥30)^28^, cancer type (obesity, non-obesity)^29^, and LTPA(insufficiently active or inactive, sufficiently active)^23^. The significance of interaction was assessed using the p-value of the product term between sleep duration and the stratified variables. Trend tests for categorical variables were conducted based on sleep duration.

Non-linear relationships were evaluated using restricted cubic splines (RCS) with 3 nodes. The analysis was adjusted for age, sex, race, PIR, educational level, BMI, smoking status, drinking status, hypertension, hyperlipidemia, diabetes mellitus, depression status, and LTPA.

Finally, sensitivity analyses were conducted, excluding deaths within the initial 2 years of follow-up to mitigate the potential impact of reverse causation.

Statistical analysis was conducted using R (version 4.4.0). A two-sided p-value less than 0.05 was considered statistically significant.

## Results

### Baseline Characteristics of Study Participants

A total of 70,190 participants were enrolled in NHANES between 2005 and 2018. Initially, individuals under 18 years of age and those lacking cancer assessment data were excluded (n = 30,476). Subsequently, participants without sleep duration data (n = 131) and pregnant individuals during the survey period (n = 709) were also excluded. Participants lacking weighting information were further excluded (n = 1,536) according to NHANES weighting guidelines. Finally, 2 participants deemed unsuitable for follow-up were excluded from survival analyses. Therefore, a total of 37,338 participants were available for logistic regression analyses, and 3,593 cancer survivors were included in survival analyses (Figure 1).

**Fig. 1:**
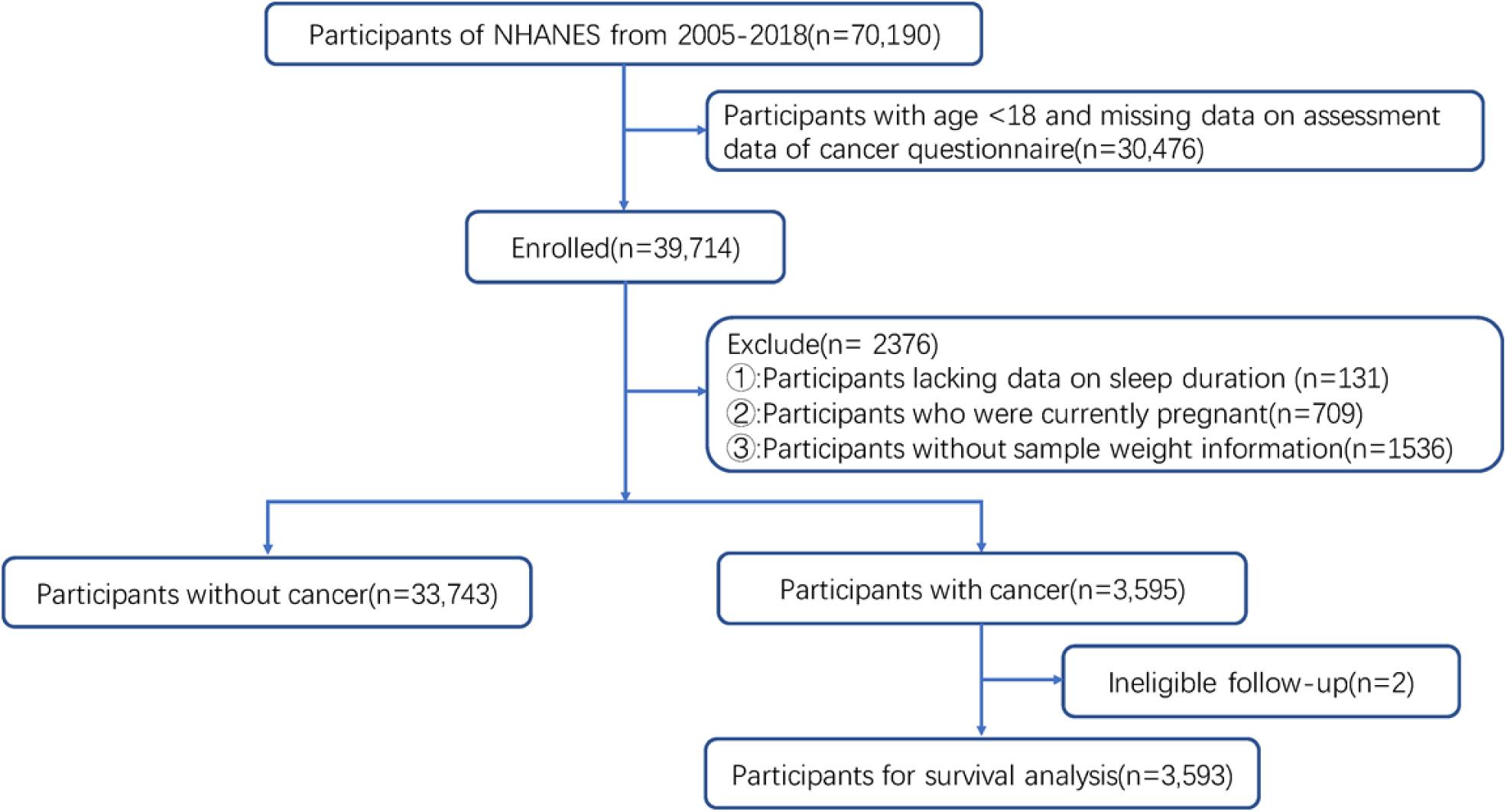
Participant flow diagram

Table 1 presents the baseline characteristics of the cancer survivor subgroup in NHANES from 2005 to 2018. The mean age of the study population was 47.59 (0.22) years, with 48.72% being male and a majority identifying as non-Hispanic white (66.85%). Overall, the weighted percentage of cancer incidence was 10.12%. Compared to non-cancer survivors, cancer survivors were more likely to be older, white females with higher levels of education and economic status. They also exhibited lower rates of smoking and alcohol use, engaged in less LTPA, and reported more hours of daily sleep.

**Table 1:**
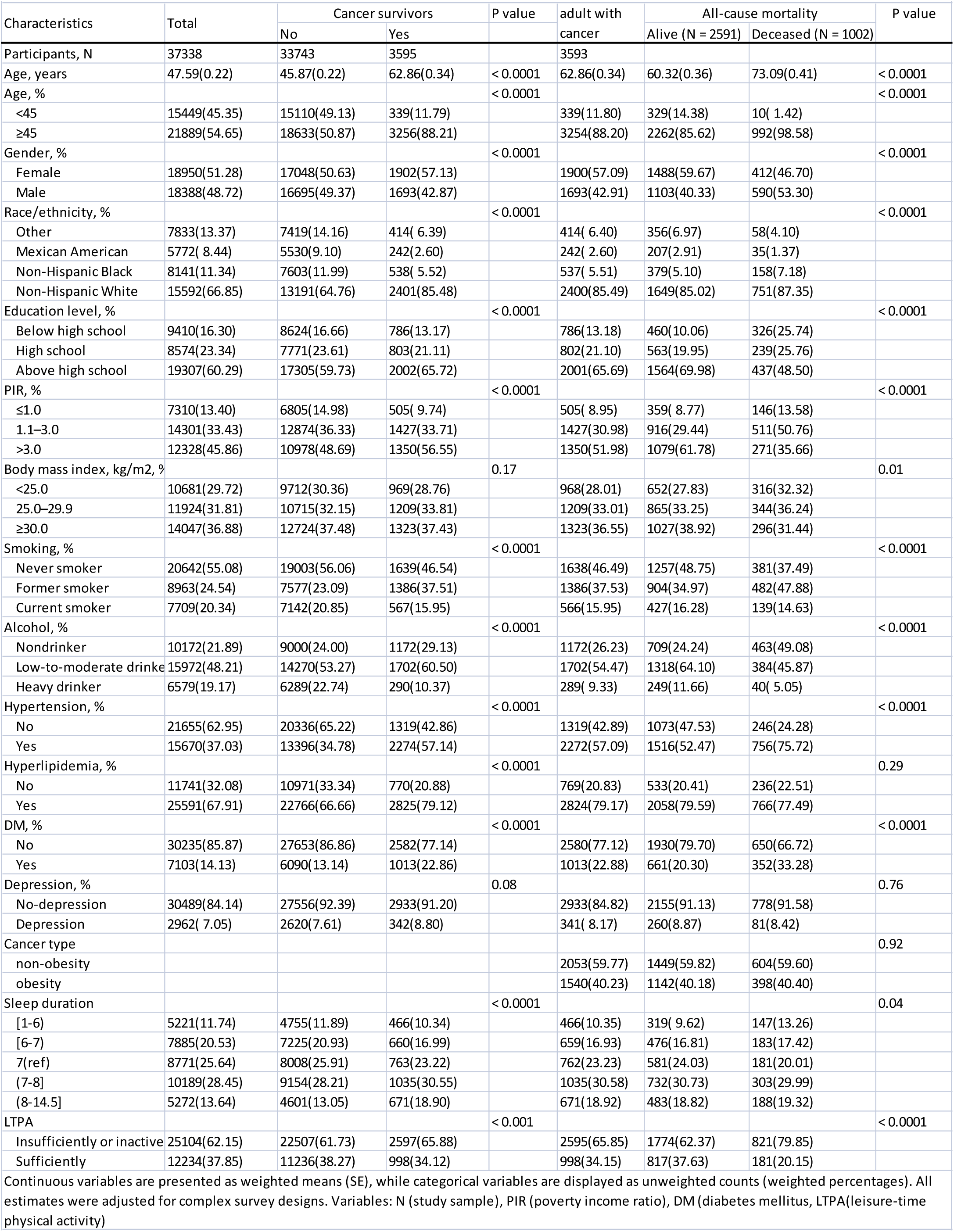
Characteristics of adult participants and adult with cancer in NHANES 2005–2018.

During a median follow-up period of 6.1 years, a total of 1002 deaths from all causes were documented, including 341 deaths related to cancer and 661 deaths from non-cancer causes (Table 1). Individuals who died from all causes were more likely to be older Hispanic white males with lower levels of education and income. They also tended to have higher BMI, engage in less LTPA, and report fewer hours of sleep compared to those who survived. Additionally, participants who died had higher rates of hypertension anddiabetes.

### Association between sleep duration and cancer incidence

Sleep duration was categorized into five groups, with 7 hours of sleep used as the reference category, and its association with cancer incidence was evaluated (Table 2). The OR and 95%CI from the original model indicated that longer sleep duration was positively associated with cancer incidence(1.21 (1.08,1.35) for (7-8] and 1.62 (1.40,1.86) for (8-14.5]). In fully adjusted multivariate regression models, the association between longer sleep durations relative to 7 hours per day and cancer incidence remained significant and consistent(1.18 (1.04,1.33) for (7-8] and 1.54 (1.28,1.86) for (8-14.5]). In all models, the trend test indicated statistically significant associations(all P _for-trend_ <0.05).

**Table 2.**
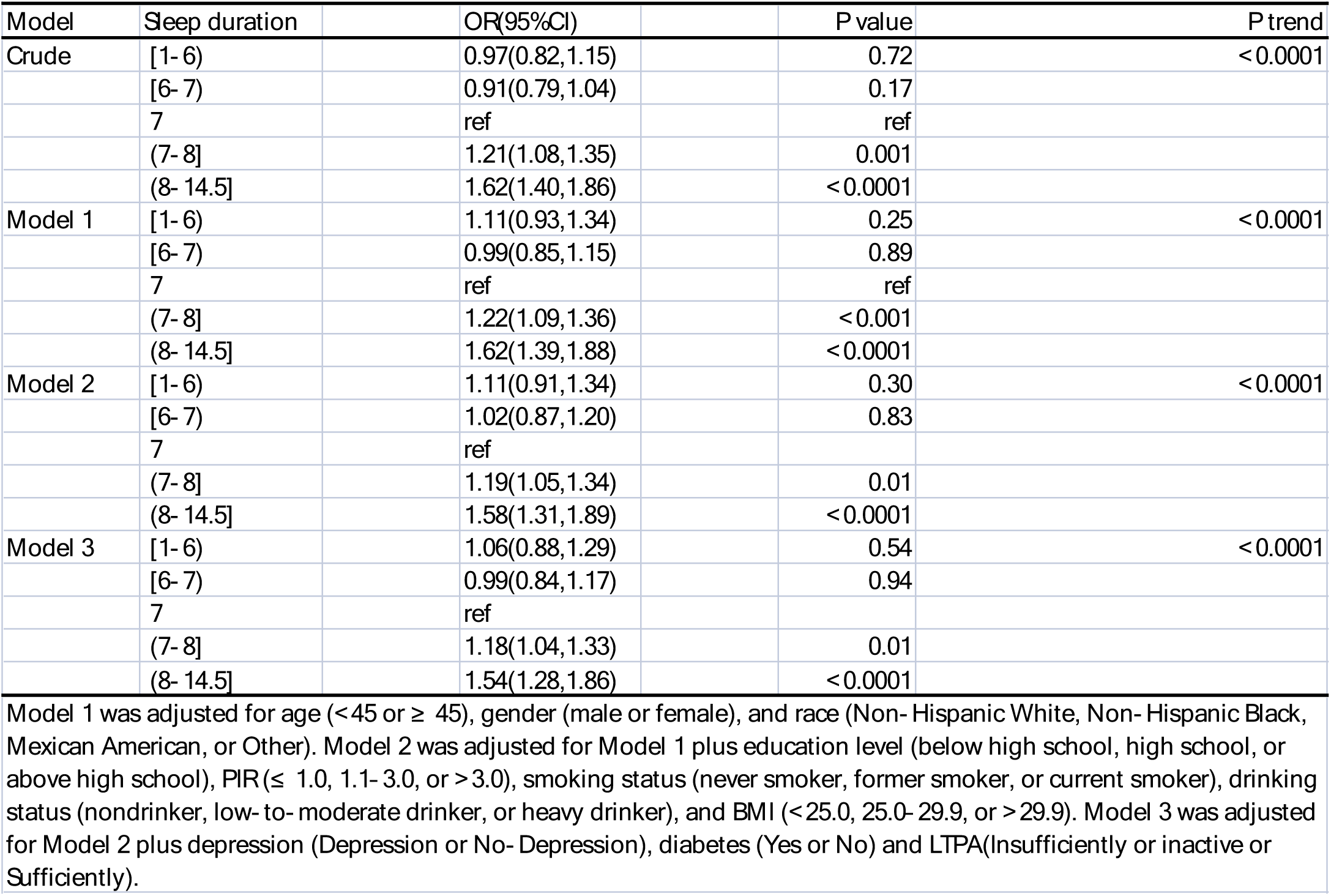
ORs (95%CIs) of the prevalence of cancer according to the Sleep duration in the NHANES2005– 2018 (n = 37,338)

Figure 2 illustrates the dose-response relationship between sleep duration and cancer incidence using RCS. The RCS analyses reveal an "L-shaped" association between sleep duration and cancer incidence in the general population. The results revealed a pronounced non-linear relationship between sleep duration and cancer incidence in the general population (Figure 2, P-overall < 0.001, P for non-linearity = 0.0003). Interestingly, within the ≥7 hours group, there was a significant positive correlation observed between sleep duration and cancer incidence (Figure 2).

**Fig. 2:**
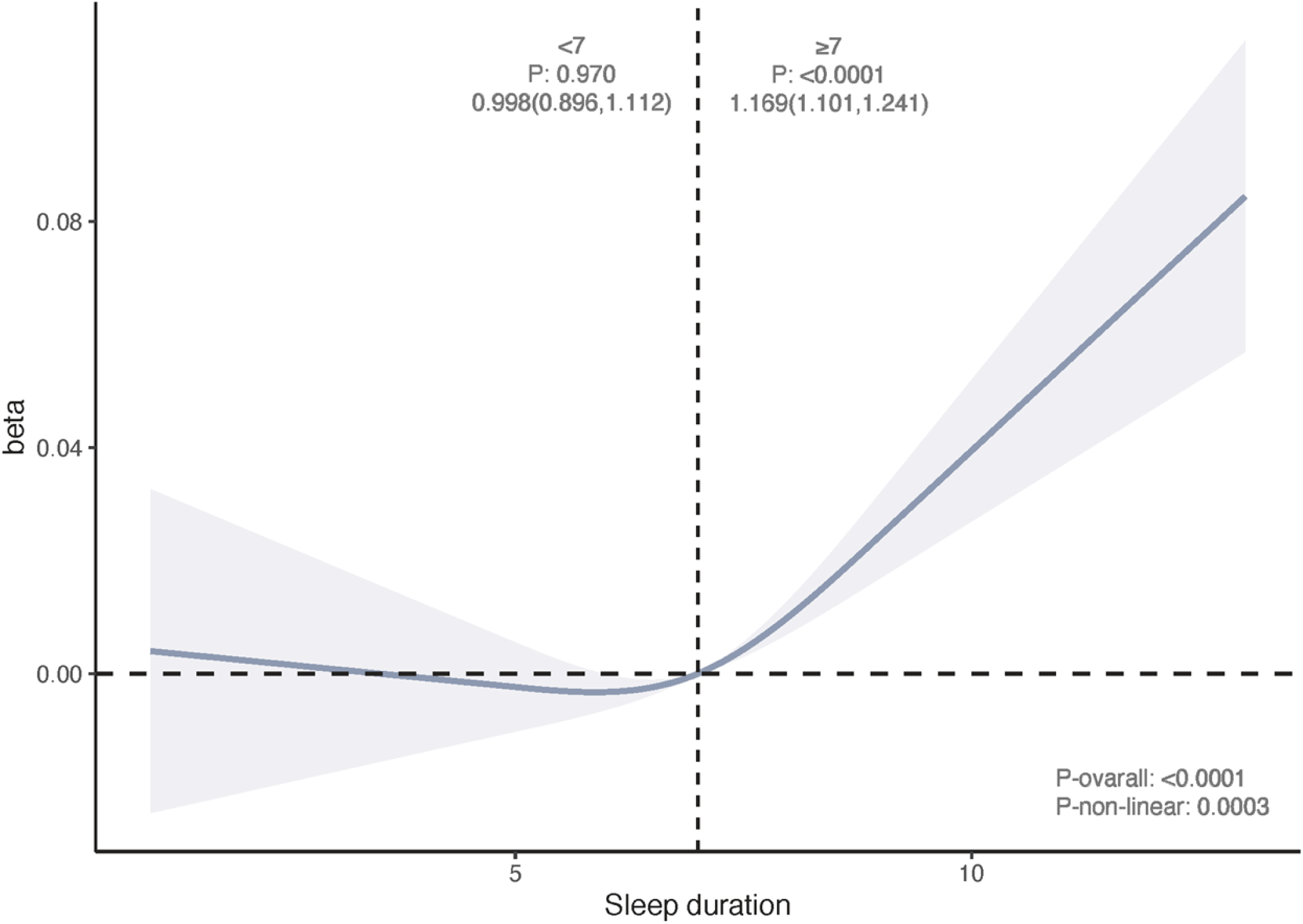
Dose-response relationship between sleep duration and cancer risk.

In subgroup analyses (Table 3), stratified by age (<45 years, ≥45 years), sex (male, female), race (white, other), BMI (<30, ≥30), and LTPA(Inactive or Insufficiently active, Sufficiently active), it was observed that among participants aged 45 years or older, females, and white participants, more than 7 hours of sleep significantly increased the risk of cancer. Importantly, the findings from the subgroup analyses were consistent with the trends observed in the primary analysis (Table 2).

**Table 3.**
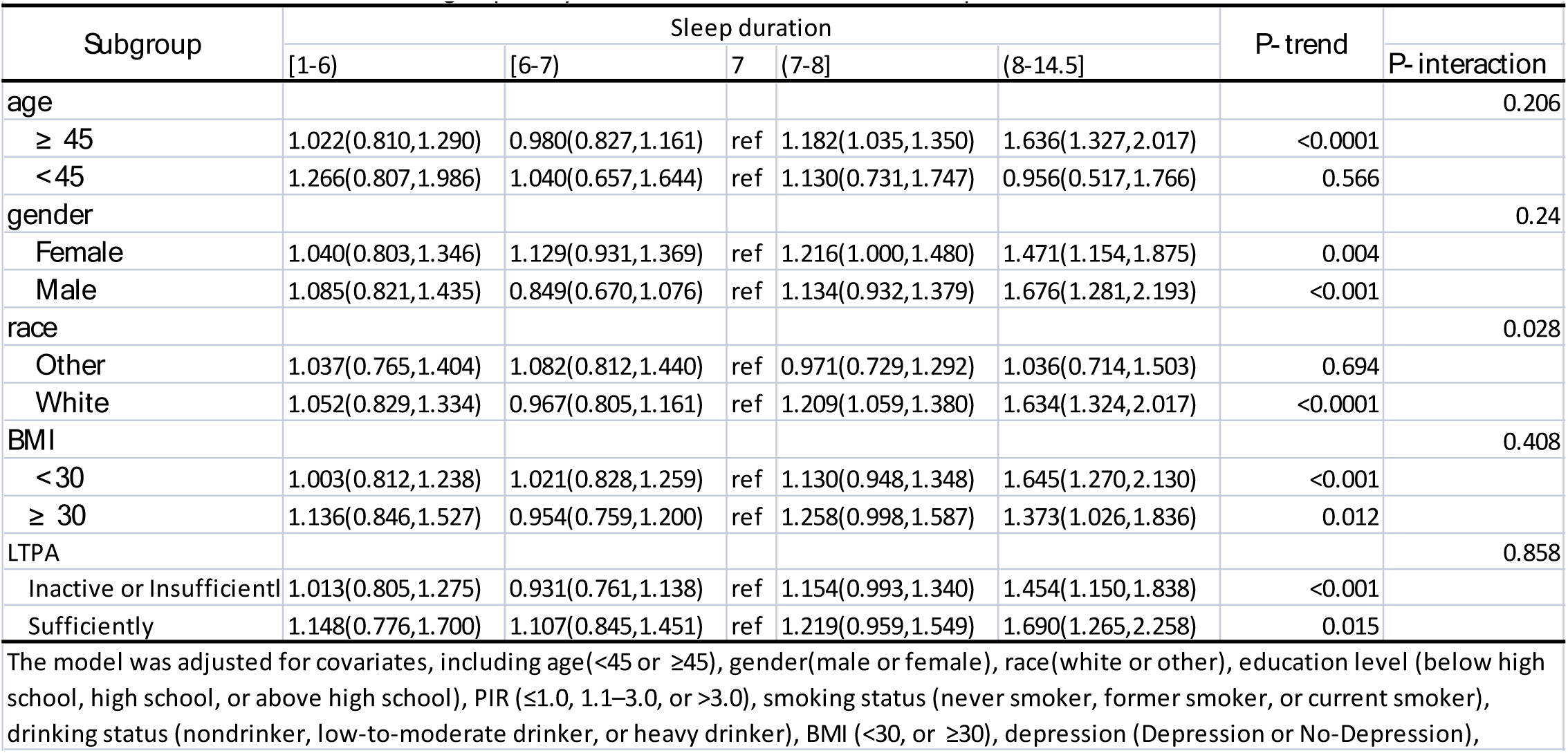
Subgroup Analysis of the Association Between Sleep Duration and Cancer Risk

### Association between sleep duration and mortality in cancer survivors

In the original model, the HR and 95%CI indicated a positive association between both longer and shorter sleep durations and all-cause and non-cancer mortality among cancer survivors. Similar trends were observed for cancer-specific mortality, although statistical significance was only reached in the longest sleep duration subgroup ((8-14.5]) (Table 4). In fully adjusted multivariate regression models, the HRs for all-cause mortality in the (7-8] and (8-14.5] sleep duration subgroups compared to 7 hours per day were 1.40 (95% CI 1.06, 1.84) and 1.90 (95% CI 1.42, 2.54), respectively. For cancer-specific mortality, the HRs in the (7-8] and (8-14.5] sleep duration subgroups were 1.20 (95% CI 0.77, 1.87) and 1.70 (95% CI 1.05, 2.73), respectively. For non-cancer-specific mortality, the HRs in the (7-8] and (8-14.5] sleep duration subgroups were 1.50 (95% CI 1.07, 2.12) and 2.03 (95% CI 1.42, 2.90), respectively.

**Table 4.**
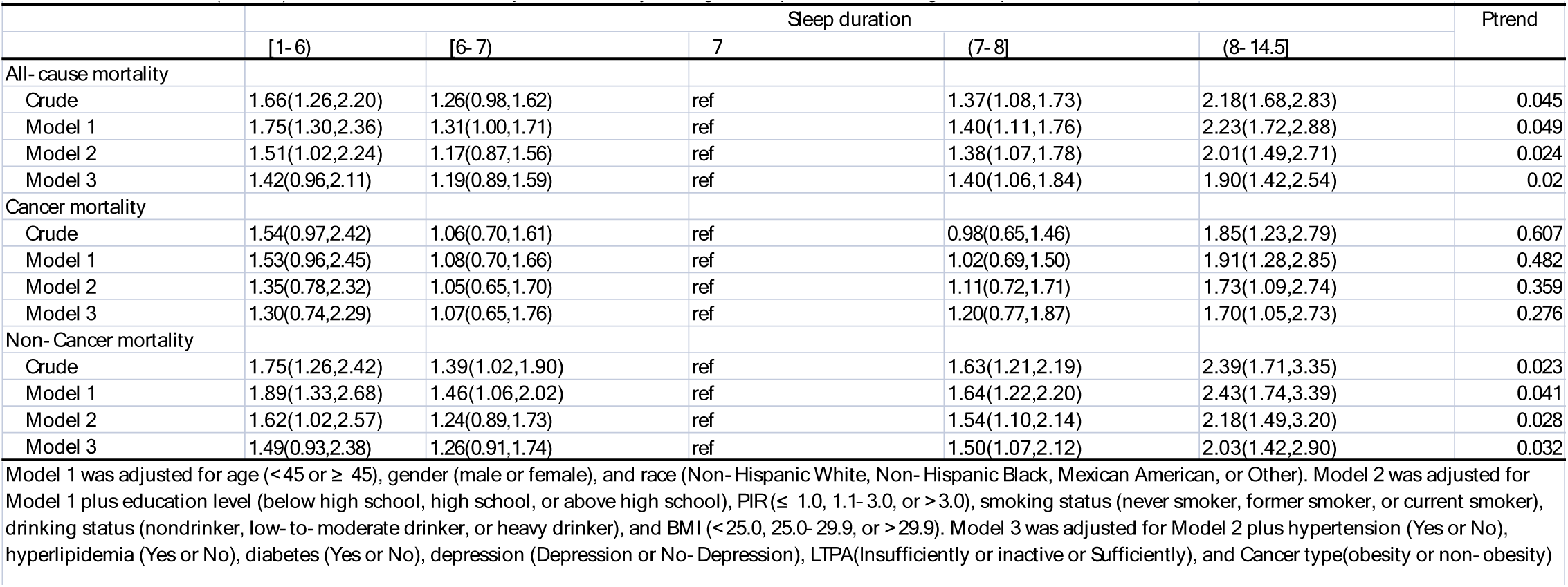
HRs (95%CIs) for all- cause and cause- specific mortality among cancer patients according to sleep duration.

The RCS plot based on survival analysis indicated a "U-shaped" association between sleep duration and mortality among cancer survivors (Figure 3). The findings highlight a robust non-linear relationship between sleep duration and mortality among community cancer survivors (all P-overall < 0.01, P for non-linearity < 0.01). Notably, the inflection point for significantly higher mortality was observed at 7 hours of sleep per day, regardless of the cause of death. Beyond this point, each additional hour of sleep was associated with a 32.4% increased risk of all-cause mortality, a 30.8% increased risk of cancer-specific mortality, and a 33.5% increased risk of non-cancer-specific mortality (Figure 3).

**Fig. 3:**
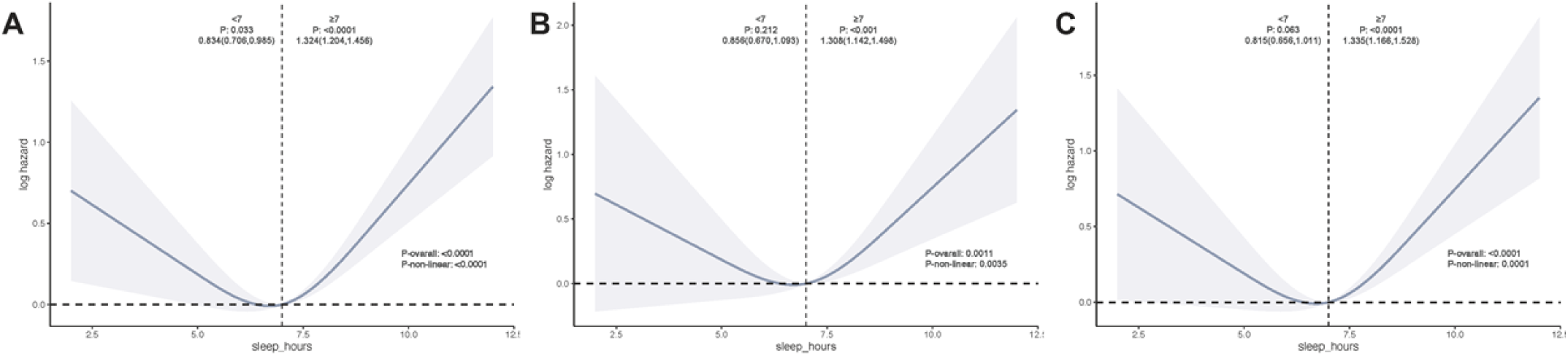
Dose-response relationship between sleep duration and risk of mortality in cancer survivors; (A) for all-cause mortality, (B) for cancer mortality, (C) for non-cancer mortality.

In subgroup analyses of all-cause mortality (Table 5), sleep durations exceeding 7 hours were found to significantly elevate the risk of mortality among survivors aged 45 years or older, males, whites, those with non-obese BMI (<30), and individuals with obesity-related cancer. Importantly, the findings from these subgroup analyses of survival were consistent with the trends observed in the primary analyses (Table 4).

**Table 5.**
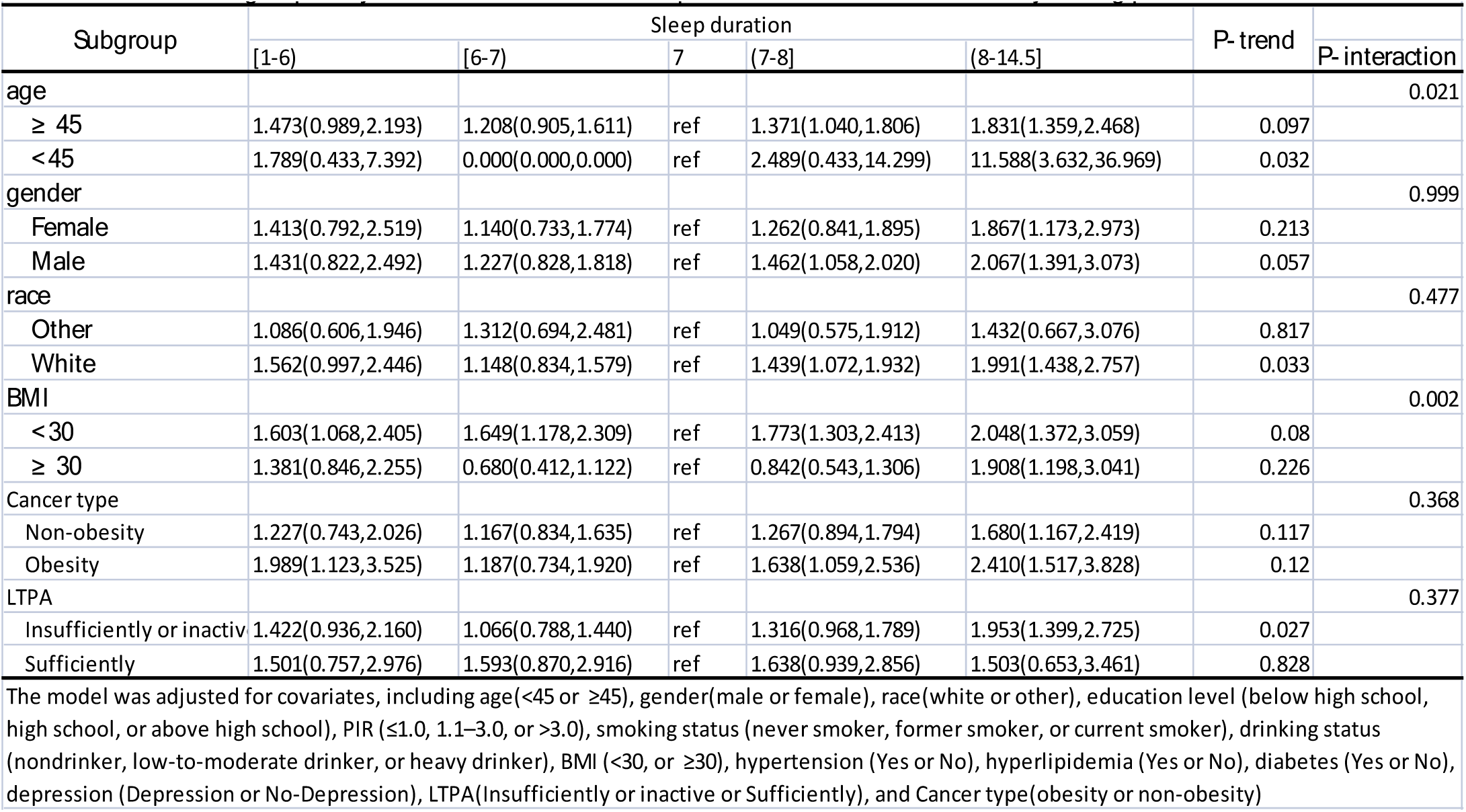
Subgroup analyses of the association of sleep duration with all- cause mortality among patients with cancer.

### Sensitivity analysis

Table S1 and Figure S1 summarize the results of the sensitivity analysis. Excluding participants who died within the first 2 years of follow-up (n = 3877), sleep durations exceeding 7 hours per day continued to show an increased risk of all-cause and non-cancer mortality (Table S1). Figure S1 depicts a similar non-linear relationship as observed in the primary analysis, with the inflection point of the curve still at 7 hours. Subgroup analyses based on this inflection point yielded results consistent with those of the primary analysis (Figure S1).

## Discussion

This study utilized data from the NHANES database and applied appropriate weighting to ensure that our analysis accurately represents the broader U.S. population. Our findings reveal that the weighted percentage of cancer survivors in the U.S. population is 10.12%. Moreover, our results indicate a positive association between longer sleep duration and increased cancer incidence in the general population. We investigated the non-linear relationship between sleep duration and cancer incidence using RCS and observed that 7 hours of sleep per day may be associated with lower cancer incidence. Importantly, there was a significant positive correlation between sleep durations exceeding 7 hours and increased cancer incidence. This study further explored the optimal sleep duration for cancer survivors. Interestingly, the results of the survival analyses consistently indicated that sleep durations exceeding 7 hours were significantly associated with all-cause, cancer-specific, and non-cancer mortality among cancer survivors. Our results are robust, as the association between sleep duration and both cancer incidence and prognosis remained stable across various analytic strategies, including adjustment for multiple confounders and sensitivity analyses. Given that sleep duration is a modifiable risk indicator influenced by personal behaviors, our study offers promising insights into its potential application in oncological diagnosis and prognostic assessment.

To our knowledge, this study represents the first comprehensive analysis examining the relationship between sleep duration and both cancer incidence and prognosis in the general U.S. population. Given the complex interplay between cancer and sleep^11,30^, investigating sleep duration’s impact on cancer progression in a nationally representative sample holds substantial public health implications. Our study demonstrated that longer sleep duration was associated with increased cancer incidence in the general population. To explore potential non-linear correlations, we conducted RCS analyses concurrently. Interestingly, the results from RCS plots aligned closely with those of our primary analyses. This consistency was observed in both the RCS plots and regression analyses based on grouped continuous variables set according to inflection points (Figure 2). We also investigated the relationship between sleep duration and prognosis among cancer survivors. Surprisingly, sleep durations exceeding 7 hours significantly increased the risk of mortality among cancer survivors. Both non-linear correlation analyses and regression analyses using grouped continuous variables based on inflection point settings yielded results consistent with our primary analysis (Table 3). Notably, in contrast to Deng et al^31^, which suggested that short sleep duration may promote tumorigenesis, our findings indicate that shorter sleep duration (<7 hours) may not be a risk factor for cancer development. Our results are consistent with two pooled analyses of cohort studies^32–34^, which suggest that racial and cancer-type differences could be contributing factors.

The relationship between sleep duration and mortality risk has long been debated. Numerous studies in the general population have indicated that both excessive and insufficient sleep durations may be strongly associated with increased mortality risk^35,36^. However, two recent systematic reviews have provided conflicting results^37,38^. Our study focused on cancer survivors, and the RCS analysis revealed a "U-shaped" curve between sleep duration and the risk of mortality in this population. This suggests that both too little and too much sleep may increase the risk of mortality. However, further stratified analyses based on the inflection point indicated that the relationship between sleep duration and mortality risk stabilized in subgroups where sleep duration was ≥7 hours(Figure 3). Our findings are supported by preclinical studies indicating that a significant portion of spontaneous tumor cell dissemination occurs during sleep. Mechanistically, it has been observed that cancer cells can be activated by melatonin during sleep, leading to increased proliferation and metastasis compared to wakeful periods^11,12,39^. Importantly, our study similarly suggests a strong association between longer sleep duration and both increased cancer incidence and poorer prognosis.

Based on the findings from our subgroup analyses, individuals aged 45 years and older who reported ≥7 hours of sleep not only showed an increased risk of cancer incidence but also demonstrated a significant elevation in mortality risk among cancer survivors. These results underscore the importance of paying closer attention to this age group in clinical and public health interventions. This phenomenon can be explained by two factors: firstly, 45 years of age is a critical point in the health trajectory of individuals, beyond which there is an increased risk of cancer and other chronic diseases^26,27^. Secondly, as individuals age, their daily sleep requirement decreases, and longer sleep durations may paradoxically increase the risk of adverse health events^6,7,37^. White females are more likely to be cancer survivors, possibly because the most common types of cancer among survivors in the United States are skin and breast cancers^18,40,41^. The observation that longer sleep increases mortality risk in males is consistent with previous findings, suggesting a stronger association between sleep characteristics and mortality in this group^42^. Additionally, we found that non-obese individuals with longer sleep durations had a higher risk of mortality compared to obese individuals, which may be related to the obesity paradox theory^43^. Finally, obesity-associated cancers may represent cancer types with a worse prognosis^23,29^.

## Strengths and limitations

Our study has several significant strengths. First, we utilized a nationally representative cohort from the United States, employing a population-based design, adjusting for a range of potential confounders, and applying the weighting strategy recommended by the NCHS. This approach allowed us to elucidate the true relationship between sleep duration and cancer incidence and mortality^25^. Second, sleep duration is an easily monitored and modifiable risk indicator, enhancing the public generalizability of our findings. Finally, at the population level, our results suggest that 7 hours of sleep may be associated with the lowest risk of cancer and mortality, providing a basis for refining recommended sleep durations.

Several limitations of this study should be noted. First, the cancer-related data relied on self-reported information from participants, which may have introduced recall bias. However, NHANES implemented standardized and rigorous control procedures to ensure data reliability and integrity. Second, despite controlling for numerous potential confounders, unmeasured variables may still affect the results. Third, due to sample size limitations, the heterogeneity of cancer-related characteristics may not be fully assessed. To mitigate this potential bias, we adjusted for cancer type in our analyses. Finally, the study data may be representative only of the U.S. population, and further generalization requires validation through global studies.

## Conclusion

In this comprehensive analysis, sleep durations of ≥7 hours were observed to significantly increase the risk of cancer development and worsen prognosis. These findings have important public health implications, suggesting that monitoring and modifying sleep behaviors may have potential applications for reducing both cancer risk and mortality among cancer survivors.

## List of abbreviations

NHANES: National Health and Nutrition Examination Survey
OR: Odds Ratio
HR: Hazard Ratio
95%CI: 95%Confidence Interval
RCS: Restricted cubic spline
CAPI: Computer Assisted Personal Interview
STROBE: Strengthening the Reporting of Observational Studies in Epidemiology
NDI: National Death Index
ICD-10: International Statistical Classification of Diseases and Related Health Problems, Tenth Revision
BMI: Body mass index
PIR: Poverty income ratio
PHQ-9: Patient Health Questionnaire-9
LTPA: Leisure-time physical activity
GPAQ: Global Physical Activity Questionnaire
NCHS: National Center for Health Statistics
SE: Standard Error

## Declarations

### Ethics approval and consent to participate

The research protocol received approval from the Institutional Review Board of the National Center for Health Statistics (NCHS), and informed consent was obtained from all participants.

### Consent for publication

Not Applicable.

### Availability of data and materials

The data used for all analyses are sourced from the public database NHANES, available at: [https://www.cdc.gov/nchs/nhanes/index.htm].

## Authors’ contributions

CQL, CZ and JS put forward ideas; CQL, CZ, JS,LL wrote the main manuscript text; CQL, CZ, JS ,JY,NC and YX, prepared figures; CQL, YX, NC and HW were used for data retrieval and statistical analysis. JMY provide funding and proofreading. All authors reviewed the manuscript.

## Data Availability

The data used for all analyses are sourced from the public database NHANES, available at:[https://www.cdc.gov/nchs/nhanes/index.htm].

## Acknowledgements

Thanks the NHANES team for the outstanding contribution.

